# Comparative Evaluation of Three Serologic Assays for the Identification of SARS-CoV-2 Antibodies

**DOI:** 10.1101/2020.08.04.20167643

**Authors:** Keenan O. Hogan, Dave Klippel, Fred V. Plapp, Rachael M. Liesman

## Abstract

**Background and aims:** Serologic assays for the detection of severe acute respiratory syndrome coronavirus 2 (SARS-CoV-2) antibodies are being developed and approved rapidly with limited external validation. Accurate diagnostics are an essential component to pandemic management and public health.

**Materials and methods:** Residual serum samples (N=113) from patients who were evaluated for SARS-CoV-2 infection status by polymerase chain reaction (PCR) were retrospectively tested in parallel across three automated SARS-CoV-2 serologic assays: Liaison SARS-CoV-2 S1/S2 IgG, Elecsys anti-SARS-CoV-2 total antibody, and Access SARS-CoV-2 IgG.

**Results:** Testing of 51 PCR-positive and 62 PCR-negative patients demonstrated qualitative inter-test agreement of 96% overall, 100% in PCR-negative patients, 88% in early positive samples (0-13 days post positive PCR), and 100% in convalescent samples (14+ days post positive PCR).

Calculated kappa values for paired inter-test agreement ranged 0.93-0.96. Compared to PCR, overall percent positive agreement ranged from 82-86% (100% for convalescent samples) and percent negative agreement was 100% for each assay.

**Conclusion:** This study demonstrates high diagnostic accuracy and inter-test agreement for three automated SARS-CoV-2 serologic assays. External validation of serologic assays is critical to ensure diagnostic accuracy and appropriate utilization of critical resources.

## 1. Introduction

Documented cases of severe acute respiratory syndrome coronavirus 2 (SARS-CoV-2) exceeded 12 million by July 12, infecting individuals from 213 countries/regions and killing more than 560,000 [1]. Management of the coronavirus disease 2019 (COVID-19) pandemic relies on accurate diagnostics, routinely provided by viral RNA detection through reverse transcriptase polymerase chain reaction (RT-PCR) and antibody detection through immunoassays. Although seroconversion has been detected as early as 4 days post symptom onset, PCR remains the current standard for identification of acute infection [2–3].

Infectious disease and microbiology organizations recommend targeted use of serologic assays for epidemiologic surveillance, identification of potential convalescent plasma donors, and evaluation of experimental vaccine efficacy [4–5]. As of the writing of this manuscript, the Food and Drug Administration (FDA) has issued emergency use authorization for 25 different serology assays [6]. These assays vary significantly in methodology, antibody targets, and manufacturer validation with few published peer-reviewed studies available for independent performance evaluation.

In this study, we assessed the performance characteristics of three automated SARS-CoV-2 serologic immunoassays using divided serum samples from patients with positive and negative SARS-CoV-2 PCR results: Liaison SARS-CoV-2 S1/S2 IgG, Elecsys anti-SARS-CoV-2 total antibody, and Access SARS-CoV-2 IgG. We hypothesized that parallel testing would demonstrate good qualitative agreement and high negative and positive percent agreement (>98%).

## 2. Material and Methods

### 2.1 Specimen Collection

Between April 15 and June 1, 2020, residual serum samples ordered for routine medical management of inpatients at the University of Kansas Hospital were retrospectively collected. Samples were collected for two groups: (1) serum samples from patients who tested positive for SARS-CoV-2 by an RT-PCR assay, and (2) serum samples from randomly selected patients who had tested negative for SARS-CoV-2 by an RT-PCR assay within 48 hours prior to collection.

SARS-CoV-2 diagnostic testing was performed using a commercially available FDA EUA RT-PCR assay. Briefly, nasopharyngeal swabs collected in either UTM or PBS were tested with the Abbott RealTime SARS-CoV-2 assay (Abbott Diagnostics Inc, Scarborough, ME), performed on the Abbott m2000 instrument, or the Simplex COVID-19 Direct assay (DiaSorin Molecular LLC, Cypress CA), following manufacturer’s instructions.

Residual serum samples were centrifuged, aliquoted, and frozen at −30 °C for 1 to 46 days. Samples were sequentially thawed and maintained at 2-8 °C for ≤ 14 days prior to testing. Patient medical records were reviewed to determine sex, age, and the date of first positive SARS-CoV-2 RT-PCR test. Samples were grouped by number of days after the first positive molecular result into 3 pre-specified categories: 0-6 days, 7-13 days, 14+ days. All available serum samples from PCR-positive patients and randomly selected PCR-negative patients that were older than 18 years with adequate residual volume for parallel testing were included.

Samples for a single PCR-positive patient from sequential days were combined when required for adequate testing volume, as long as the combined samples fell within the same pre-specified category. Clinical Laboratory Scientists were not specifically blinded to the clinical status or PCR results of the patients. This study was approved by the University of Kansas Medical Center institutional review board prior to testing.

### 2.2 Serologic Assays

Table 1 provides a breakdown of key attributes for each serologic assay. The Elecsys anti-SARS-CoV-2 antibody test (Roche Diagnostics, Rotkreuz, Switzerland) is a double antigen sandwich electro-chemiluminescence immunoassay that detects total antibody against the nucleocapsid (N) antigen and is performed on the Cobas e801 immunoassay analyzer. Testing was performed in accordance with manufacturer’s instructions. The patient sample is incubated with biotinylated SARS⍰CoV⍰2⍰ recombinant N antigen labeled with a ruthenium complex. Streptavidin-coated magnetic microparticles are added and the antigen-antibody-antigen complex binds to the solid phase by interaction of biotin with streptavidin. Microparticles are magnetically captured onto the surface of an electrode where application of a voltage induces chemiluminescence which is measured by a photomultiplier. Results are expressed as a cut-off index (COI). A result of <1.0 is considered non-reactive while a result of ≥1.0 is considered reactive.

**TABLE 1.** Attributes for the evaluated serologic assaysAbbr.

|  | Roche | DiaSorin | Beckman |
| --- | --- | --- | --- |
| Assay method | Electro CLIA, Total Ab | Indirect CLIA, IgG | CLIA, IgG |
| Antigen | Nucleocapsid protein | S1/S2 protein | S1 protein RBD |
| Specimen type | Serum, Plasma | Serum, Plasma | Serum, Plasma |
| Total volume | 250 μL | 170 μL | 250 μL |
| Result units | Cutoff Index (COI) | Arbitrary Unit (AU)/mL | Signal/Cutoff (S/CO) |
| Positive cutoff | $\geq 1.0$ COI | $\geq 15.0$ AU/mL | $\geq 1.0$ S/CO |
| Equivocal cutoff | None | None | $\geq 0.8$ and $< 1.0$ S/CO |
| Throughput | 300 samples per hour | 170 samples per hour | 400 samples per hour |
| Abbr.: Ab, Antibody; CLIA, Chemiluminescent Immunoassay; RBD, Receptor Binding Domain |  |  |  |

The Liaison SARS-CoV-2 S1/S2 assay (DiaSorin S.p.A., Saluggia, Italy) detects IgG antibody against the S1 and S2 subunits of the spike protein using a chemiluminescent indirect immunoassay (CLIA) and is performed on the DiaSorin Liaison XL immunoassay analyzer.

Testing was performed in accordance with manufacturer’s instructions. Patient serum is combined with magnetic particles coated with biotinylated recombinant S1/S2 antigen, which binds to SARS-CoV-2 antibody, if present. Following incubation, isoluminol labeled mouse anti-human IgG are added, which will bind to the SARS-CoV-2 antibody. The amount of isoluminol antibody conjugate is determined as a flash chemiluminescence reaction and reported in arbitrary units per milliliter (AU/mL). A result of <15 is considered negative while a result of ≥15.0 is considered positive.

The Access SARS-CoV-2 IgG assay (Beckman Coulter, Inc., Minnesota, USA) is a chemiluminescent indirect immunoassay that detects IgG antibody to SARS-CoV-2 and is performed on the Beckman Coulter DXI 800 immunoassay analyzer. Testing was performed in accordance with manufacturer’s instructions. A sample is incubated with paramagnetic particles coated with recombinant SARS-CoV-2 protein specific for the receptor binding domain (RBD) of the S1 Protein. Monoclonal anti-human IgG alkaline phosphatase conjugate is added to bind IgG antibodies captured on the particles. Light generated by chemiluminescent substrate is measured with a luminometer. The light signal is compared to the cut-off value and is expressed as a signal to cutoff ratio (S/CO). A result of <0.8 is interpreted as non-reactive while a result of ≥1.0 is considered reactive. Results between 0.8 and 1.0 (inclusive) are considered equivocal.

### 2.3 Statistics

Each sample represented a unique patient. For the purposes of analysis, equivocal results were treated as negative and combined samples were represented by the day farthest from the patient’s positive PCR test. Percent positive agreement (PPA) and percent negative agreement (PNA) were calculated with 95% confidence intervals (CI) using the Clopper-Pearson exact method for sensitivity and specificity and stratified by length of time from positive PCR test. Paired inter-test agreement was calculated with 95% CI and standard error (SE) using Fleiss kappa. Level of agreement was pre-specified as almost perfect (0.81-0.99), substantial (0.61-0.80), moderate (0.41-0.60), fair (0.21-0.40), and slight (0.00-0.20). Statistical analysis was performed using GraphPad Prism 8 statistics software (San Diego, USA).

## 3. Results

### 3.1 Patient Demographics and Specimen Properties

Fifty-one specimens from unique PCR-positive patients and 62 specimens from unique PCR-negative patients were available for antibody testing. Table 2 describes patient demographics stratified by PCR status. Patients were 60% female with a median age of 59 years (range: 19-92). Date of symptom onset was not routinely documented so patients were grouped temporally by the period between the first positive PCR result and serology specimen collection. Each temporal group contained 17 specimens. The number of days post positive PCR ranged from 1-45 days overall (median: 9) with a median of 5, 9, and 18 for 0-6 days, 7-13 days, and 14+ days, respectively. Samples were combined for 14 patients 0-6 days, 3 patients 7-13 days, and 2 patients 14+ days.

**TABLE 2.** Patient demographics by PCR status.

|  | Total | PCR (+) |  |  | PCR (-) |
| --- | --- | --- | --- | --- | --- |
|  |  | 0-6 days | 7-13 days | 14+ days | Total |
| Number of patients, n | 113 | 17 | 17 | 17 | 62 |
| Median age in years (IQR) | 59 (43-72) | 71 (52-77) | 64 (42-74) | 64 (55-69) | 53 (35-70) |
| Female, n (%) | 69 (61) | 12 (71) | 9 (53) | 9 (53) | 39 (63) |

### 3.2 Diagnostic Accuracy

Of 113 patients, there was consensus between the platforms in 96% (41 positive and 68 negative). There was 100% consensus in the convalescent positive samples (14+ days post positive PCR) and negative samples but only 88% consensus in early positive samples (0-13 days post positive PCR). The only equivocal result was returned by the Beckman assay on a single non-consensus sample. Table 3 details the results of each assay compared to PCR with diagnostic accuracy and estimates of precision. No false positive results were detected and the PNA was calculated at 100% for each platform. Convalescent positive samples also demonstrated zero false negative results for a calculated 100% PPA. The PPA ranged from 59-94% for the early positive samples.

**TABLE 3.** Performance characteristics of each platform by PCR status.

|  | PCR (+), n |  |  |  | PCR (-), n |
| --- | --- | --- | --- | --- | --- |
|  | 0-6 days | 7-13 days | 14+ days | Total | Total |
| Roche |  |  |  |  |  |
| Reactive | 10 | 16 | 17 | 43 | 0 |
| Non-reactive | 7 | 1 | 0 | 8 | 62 |
| PPA, % (95% CI) | 59 (33-82) | 94 (71-99) | 100 (80-100) | 84 (71-93) |  |
| PNA, % (95% CI) | 100 (94-100) | 100 (94-100) | 100 (94-100) | 100 (94-100) |  |
| DiaSorin |  |  |  |  |  |
| Positive | 10 | 15 | 17 | 42 | 0 |
| Negative | 7 | 2 | 0 | 9 | 62 |
| PPA, % (95% CI) | 59 (33-82) | 88 (64-99) | 100 (80-100) | 82 (69-92) |  |
| PNA, % (95% CI) | 100 (94-100) | 100 (94-100) | 100 (94-100) | 100 (94-100) |  |
| Beckman |  |  |  |  |  |
| Reactive | 11 | 16 | 17 | 44 | 0 |
| Non-reactive | 6<sup>a</sup> | 1 | 0 | 7 | 62 |
| PPA, % (95% CI) | 65 (38-86) | 94 (71-99) | 100 (80-100) | 86 (74-94) |  |
| PNA, % (95% CI) | 100 (94-100) | 100 (94-100) | 100 (94-100) | 100 (94-100) |  |

Abbr.: PPA, percent positive agreement; PNA, percent negative agreement
<sup>a</sup> Includes 1 equivocal result

Two patient specimens were reported as negative by at least 1 platform in the 7-13 days post positive PCR group and both specimens were collected at 7 days. One tested negative by all three platforms and review of the medical record revealed the patient was receiving immunosuppressive therapy following renal transplantation. The second patient tested positive on the Roche and Beckman platforms and negative on the DiaSorin platform. This patient was moderately symptomatic with an elevated lactate and required supplemental oxygen but not intubation during their disease course. This result may have represented early seroconversion as the patient was 8 days post symptom onset with low-positive reactivity for the Roche and Beckman assays (14.1 COI and 11.9 S/CO, respectively) and high-negative reactivity for the DiaSorin assay (11.3 AU/mL).

Eight patient samples collected 0-6 days post positive PCR demonstrated at least one negative serology result. The samples ranged from 1-6 days post positive PCR (median: 4). Out of the 8 samples, 5 were consensus negative, 1 was reactive by Roche alone, 1 was reactive by Beckman alone, and 1 was reactive by DiaSorin and Beckman.

### 3.3 Assay Comparison

Direct comparison of the three assays is detailed in Table 4. Calculated kappa values for inter-test agreement between Roche/DiaSorin, Roche/Beckman, and DiaSorin/Beckman were 0.94, 0.93, and 0.96, respectively. The confidence interval for each measure was within the bounds of near perfect agreement and included perfect agreement (1.00).

**TABLE 4.** Serologic assay comparison.

|  |  | DiaSorin (n) |  | Beckman (n) |  |
| --- | --- | --- | --- | --- | --- |
|  |  | (+) | (-) | (+) | (-) |
| Roche (n) | (+) | 41 | 2 | 42 | 1 |
|  | (-) | 1 | 69 | 3 | 68 |
|  | K<sup>a</sup> | 0.94<br>(0.88-1.0, 0.032) |  | 0.93<br>(0.86-1.0, 0.036) |  |
| Beckman (n) | (+) | 42 | 2 |  |  |
|  | (-) | 0 | 69 |  |  |
|  | K<sup>a</sup> | 0.96<br>(0.91-1.0, 0.026) |  |  |  |

<sup>a</sup> Fleiss kappa (95% CI, SE)

## 4. Discussion

### 4.1 Comparative Analysis

This study describes high PNA and temporally dependent PPA for three automated SARS-CoV-2 serologic assays in an independent evaluation using an inpatient cohort verified by PCR. The results of this study confirm the conclusions of the systematic review by La Marca, et al. in which they suggest the choice of serology assay should be made by individual labs based on experience with the manufacturer, expected workflow and volume, and thoroughness of assay guidance and validation [7]. They report on 49 serology studies primarily conducted in China with a range of 78-100% and 0-100% for specificity and sensitivity, respectively.

Two peer-reviewed articles and three studies released as pre-prints specifically reported on the accuracy of the DiaSorin SARS-CoV-2 serologic assay in comparison to PCR, with a range of 96.8-100% for specificity and 77.8-95.7% for sensitivity in convalescent patients [8–12]. With 100% PNA and 100% PPA in the convalescent group, this study describes a more favorable diagnostic accuracy. One of the studies provided the median positive quantitative results for comparison, which ranged from 8-96.3 AU/mL, with 96.3 AU/mL representing their most acutely ill subjects [11]. In comparison, the median positive result in this study was 190 AU/mL, which may suggest our high PPA is a consequence of high patient acuity. Correlation between disease severity and antibody titer and detection has been reported [13–14]. However, direct comparison of the quantitative results from these qualitative assays is limited in significance, especially given the heterogeneity in study design and reporting as identified in a recent Cochrane review [15].

Median negative results were comparable across the studies (below the limit of detection to 5.7 AU/mL), but two of the studies included pre-pandemic cohort samples of greater than 1000 patients. The difference in sampling likely explains the discordance in reported specificity.

Additionally, two of these studies and an additional study released as a pre-print report on the Roche assay with a range of 98-99.7% and 89.2-100% for specificity and sensitivity, respectively, and similar comparisons to this study may be drawn [10,12,16].

To our knowledge, no study has reported independent evaluation of the Beckman assay diagnostic accuracy. With kappa values of 0.93 and 0.96 in comparison to the Roche and DiaSorin assay, respectively, 100% PNA, and 100% PPA in convalescent specimens, we demonstrate no significant difference in the performance between these three platforms. Including several weak positive results, seroconversion was detected across all PCR-positive patients by day 8 as shown in Figure 1. Although the time of symptom onset for our patient population was not routinely recorded, our data likely coincides with the reported median seroconversion for total antibody and IgG antibody alone of 11 and 14 days post symptom onset, respectively [17]. Although the Roche assay detects total antibody and DiaSorin and Beckman assays detect IgG antibody, there was no identifiable difference in the detection of early seroconversion.

**FIGURE 1.**
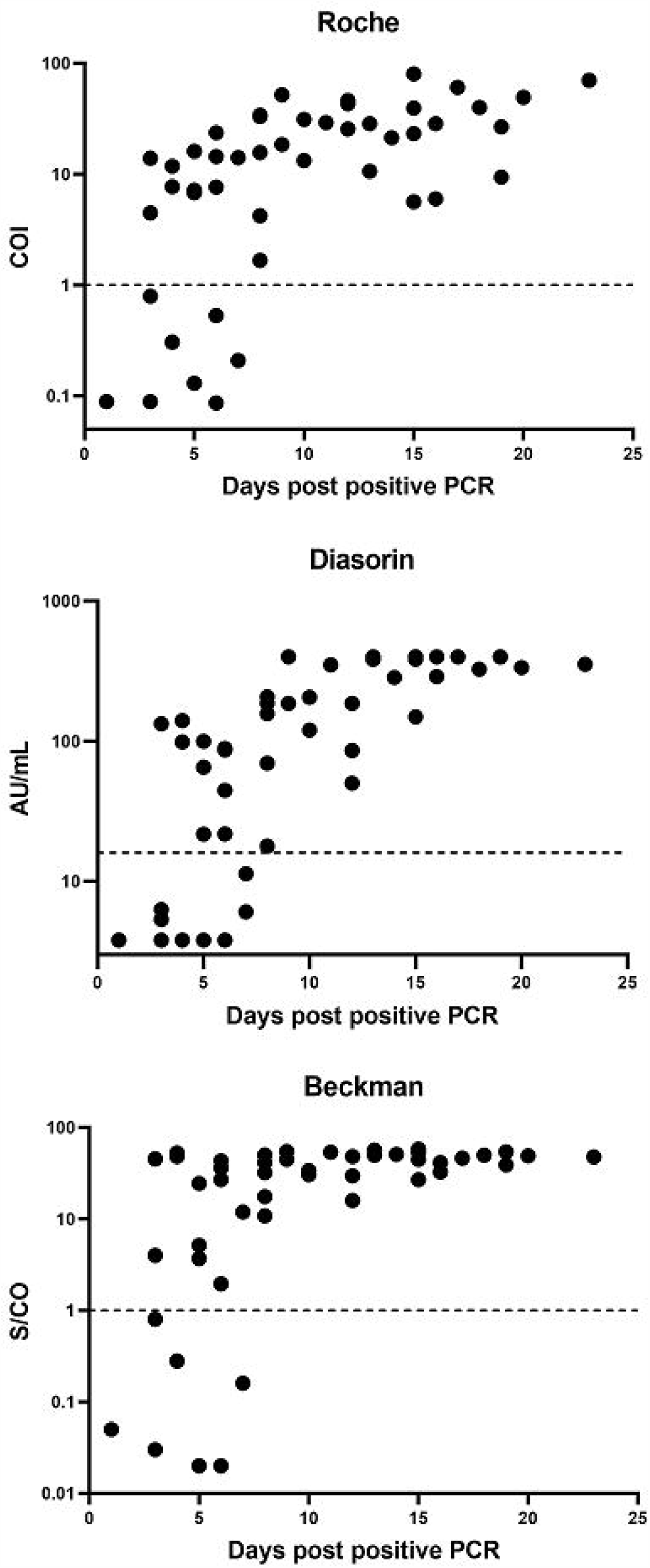
Quantitative results for each assay by days post positive PCR.

Clinical context will be essential in identifying pre-test probabilities based on local seroprevalence, patient symptomatology, and stage of disease course [18]. Although our population did not include any false positive results, testing of large pre-pandemic cohort samples have reported variable specificity. The use and interpretation of serologic assays is also complicated by the continued uncertainty of antibody durability and the relationship between positive serologic testing and the presence of neutralizing antibodies [14]. One study comparing the DiaSorin assay to a viral microneutralization test reported a sensitivity and specificity of 43.8% and 94.9%, respectively, for 70 PCR-positive patients and 81 pre-pandemic patients [19]. Overall, evidence is limited and laboratories offering serology testing for SARS-CoV-2 must make these limitations clear. For patients with a complicated medical history or atypical presentation, antibody testing may provide important diagnostic information. It has been reported that overall sensitivity for serology is greater than nucleic acid testing for SARS-CoV-2 starting day 8 after symptom onset and combined serology and RT-PCR testing provides significantly improved sensitivity compared to RT-PCR alone early in the disease course [17,20].

### 4.2 Limitations

This study has several limitations. The testing of a relatively small number of retrospective convenience samples is at risk for confirmation basis and underpowered calculations. Specifically, our cohort of inpatients likely includes a greater average patient acuity, which would tend to inflate sensitivity. This may also increase the number of false positive results, which was not identified. In the absence of thorough chart review for the PCR-negative cohort, the presence of confounding factors contributing to an artificially high true negative rate cannot be excluded but is considered unlikely. By combining specimens from sequential days and using the farthest date, we likely reduced the reportable sensitivity. Longitudinal samples from individual patients were not tested to track seroconversion. The samples were not tested to ascertain the presence of potential cross-reactive viruses.

One of the strengths of this study is the direct comparison of the three assays by parallel testing, including an assay without currently published external validation. Each sample was collected from a single patient, which avoids bias introduced by multiple measurements. Although our specificity population was smaller, the patient samples are representative of the current local circulating viruses among individuals with healthcare contacts, which lends external validity.

The results also include early seroconversion specimens in addition to convalescent specimens. The performance characteristics of serologic assays for patients in this window period is critical for clinical management decisions, particularly for acutely ill patients. We reported no false positive results that may have otherwise been attributed to remote exposure with the use of a post-pandemic specificity cohort.

### 4.3 Conclusion

We report high diagnostic accuracy and inter-test agreement for three SARS-CoV-2 serologic assays. The choice of assay and its clinical utility will depend on the specific needs of individual laboratories and the local public health conditions. External validation of serologic assays is critical to ensure high diagnostic accuracy and appropriate utilization of critical resources.

## Data Availability

No supplementary material or external datasets are available for this article.

## Funding

This research did not receive any specific grant from funding agencies in the public, commercial, or not-for-profit sectors.

## Declarations of Interest

None.

## Notes

### Competing Interest Statement

The authors have declared no competing interest.

### Author Declarations

University of Kansas Medical Center Human Research Protection Program Institutional Review Board Study #00145715

## References

[1] COVID-19 Dashboard by the Center for Systems Science and Engineering (CSSE) at Johns Hopkins University. 2020. https://coronavirus.jhu.edu/map.html. [Accessed July 12, 2020].

[2] Sethuraman N, Jeremiah SS, Ryo A. Interpreting Diagnostic Tests for SARS-CoV-2. JAMA. 2020;323(22):2249–2251. doi:10.1001/jama.2020.8259

[3] Xiang F, Wang X,1 He X, Peng Z, Yang B, Zhang J, Zhou Q, Ye H, Ma Y, Li H, Wei X, Cai P, Ma W. Antibody Detection and Dynamic Characteristics in Patients with COVID-19. Clin Infect Dis. 2020 Apr 19. doi: 10.1093/cid/ciaa461

[4] Infectious Disease Society of America. COVID-19 Antibody Testing Primer. May 2020. https://www.idsociety.org/globalassets/idsa/public-health/covid-19/idsa-covid-19-antibody-testing-primer.pdf. [Accessed July 12, 2020].

[5] Patel R, Babady E, Theel ES, Storch GA, Pinsky BA, George KS, Smith TC, Bertuzzi S. Report from the American Society for Microbiology COVID-19 International Summit, 23 March 2020: Value of Diagnostic Testing for SARS–CoV-2/COVID-19. mBio, March 2020, 11(2), [e00722–20]. https://doi.org/10.1128/mBio.00722-20

[6] Food and Drug Administration. In Vitro Diagnostics EUAs. https://www.fda.gov/medical-devices/coronavirus-disease-2019-covid-19-emergency-use-authorizations-medical-devices/vitro-diagnostics-euas. [Accessed June 28, 2020].

[7] La Marca A, Capuzzo M, Paglia T, Roli L, Trenti T, Nelson SM. Testing for SARS-CoV-2 (COVID-19): a systematic review and clinical guide to molecular and serological in-vitro diagnostic assays. Reprod Biomed Online. 2020 Jun 14. doi: 10.1016/j.rbmo.2020.06.001

[8] Tré-Hardy M, Wilmet A, Beukinga I, Dogné J-M, Douxfils J, Blairon L. Validation of a chemiluminescent assay for specific SARS-CoV-2 antibody. Clinical Chemical Laboratory Medicine. 2020 May 25. doi: 10.1515/cclm-2020-0594.

[9] Plebani M, Padoan A, Negrini D, Carpinteri B, Sciacovelli L. Diagnostic performances and thresholds: The key to harmonization in serological SARS-CoV-2 assays? Clin Chim Acta. 2020 Oct;509:1–7. doi: 10.1016/j.cca.2020.05.050

[10] Ekelund O, Ekblom K, Somajo S, Pattison-Granberg J, Olsson K, Petersson A. High-throughput immunoassays for SARS-CoV-2, considerable differences in performance when comparing three methods. medRxiv. May 26, 2020. doi: https://doi.org/10.1101/2020.05.22.20106294

[11] Bonelli F, Sarasini A, Zierold C, Calleri M, Bonetti A, Vismara C, et al. Clinical And Analytical Performance Of An Automated Serological Test That Identifies S1/S2 Neutralizing IgG In Covid-19 Patients Semiquantitatively. bioRxiv. 2020;25:2000082–35.

[12] Perkmann T, Perkmann-Nagele N, Breyer MK, Breyer-Kohansal R, Burghuber OC, Hartl S, Aletaha D, Sieghart D, Quehenberger P, Marculescu R, Mucher P, Strassl R, Wagner OF, Binder CJ, Haslacher H. Side by side comparison of three fully automated 1 SARS-CoV-2 antibody assays with a focus on specificity. medRxiv. June 9, 2020. doi: https://doi.org/10.1101/2020.06.04.20117911

[13] Yongchen Z, Shen H, Wang X, Shi X, Li Y, Yan J, Chen Y, Gu B. Different Longitudinal Patterns of Nucleic Acid and Serology Testing Results Based on Disease Severity of COVID-19 Patients Emerg Microbes Infect. 2020 Dec;9(1):833–836. doi: 10.1080/22221751.2020.1756699.

[14] Long Q, Tang X, Shi Q, Li Q, Deng H, Yuan J, Hu J, Xu W, Zhang Y, Lv F, Su K, Zhang F, Gong J, Wu B, Liu X, Li J, Qiu J, Chen J, Huang A. Clinical and immunological assessment of asymptomatic SARS-CoV-2 infections. Nat Med (2020). https://doi.org/10.1038/s41591-020-0965-6

[15] Deeks JJ, Dinnes J, Takwoingi Y, Davenport C, Spijker R, Taylor-Phillips S, Adriano A, Beese S, Dretzke J, Ferrante di Ruffano L, Harris IM, Price MJ, Dittrich S, Emperador D, Hooft L, Leeflang MMG, Van den Bruel A. Antibody tests for identification of current and past infection with SARS_JCoV_J2. Cochrane Database Syst Rev 2020 Jun 25;6:CD013652. doi: 10.1002/14651858.CD013652

[16] Tang MS, Hock KG, Logsdon NM, Hayes JE, Gronowski AM, Anderson NW, Farnsworth CW. Clinical Performance of the Roche SARS-CoV-2 Serologic Assay. Clin Chem. 2020 Jun 2:hvaa132. doi: 10.1093/clinchem/hvaa132

[17] Zhao R, Li M, Song H, Chen J, Ren W, Feng Y, Gao GF, Song J, Peng Y, Su B, Guo X, Wang Y, Chen J, Li J, Sun H, Bai Z, Cao W, Zhu J, Zhang Q, Sun Y, Sun S, Mao X, Su J, Chen X, He A, Gao W, Jin R, Jiang Y, Sun L. Early detection of SARS-CoV-2 antibodies in COVID-19 patients as a serologic marker of infection. Clin Infect Dis. 2020. doi: 10.1093/cid/ciaa523.May1pii:ciaa523

[18] Cheng MP, Yansouni CP, Basta NE, Desjardins M, Kanjilal S, Paquette K, Caya C, Semret M, Quach C, Libman M, Mazzola L, Sacks JA, Dittrich S, Papenburg J. Serodiagnostics for Severe Acute Respiratory Syndrome–Related Coronavirus-2: A Narrative Review. Ann Intern Med. 2020 Jun 4. M20-2854. doi: 10.7326/M20-2854

[19] Jaaskelainen AJ, Kuivanen S, Kekalainen E, Ahava MJ, Loginov R, Kallio-Kokko H, Vapalahti O, Jarva H, Kurkela S, Lappalainen M. Performance of six SARS-CoV-2 immunoassays in comparison with microneutralisation. J Clin Virol. 2020 Aug;129:104512. doi: 10.1016/j.jcv.2020.104512

[20] Espejo AP, Akgun Y, Al Mana AF, Tjendra Y, Millan NC, Gomez-Fernandez C, Cray C. Review of Current Advances in Serologic Testing for COVID-19. Am J Clin Pathol. 2020 June 25;aqaa112. doi: 10.1093/ajcp/aqaa112

